# Maternal health care services utilization in the amid of COVID-19 pandemic in West Shoa Zone, Central Ethiopia

**DOI:** 10.1101/2020.10.09.20210054

**Authors:** Kababa Temesgen, Negash Wakgari, Bikila Tefera, Belay Tafa, Getu Alemu, Fekadu Wandimu, Tolera Gudisa, Tolosa Gishile, Gurmessa Daba, Gizachew Abdissa, Bikila Soboka

## Abstract

The novel coronavirus (COVID-19) is an infectious disease caused by a newly discovered coronavirus. Despite strong efforts that have been taking place to control the pandemic globally, the virus is on the rise in many countries. Hence, this study assessed the maternal health care services utilization in the amid of the COVID-19 pandemic in West Shoa Zone, Central Ethiopia. Community-based cross-sectional study was conducted among 844 pregnant women or those gave birth in the last 6 months before the study. A multi-stage sampling technique was used to select the study participants. The data were collected through face-to-face interview using a semi-structured questionnaire. Logistic regressions were performed to identify the presence of significant associations, and adjusted odds ratio with 95%CI was employed for the strength and directions of association between the independent and outcome variables. A P-value of <0.05 was used to declare statistical significance. The prevalence of maternal health service utilization during the COVID-19 pandemic was 64.8%. The odds of maternal health service utilization was higher among mothers who had primary (AOR=2.16, 95%CI: 1.29-3.60), secondary (AOR=1.97, 95%CI: 1.13-3.44), and college and above education (AOR=2.89, 95%CI: 1.34-6.22) than those who could not read and write. In addition, mothers who did travel 25-74 km (AOR= 0.37, 95%CI: 0.23-0.59) and 75-99 km (AOR= 0.10, 95%CI: 0.05-0.19) to reach health facility had a lower odds of maternal health service utilization than those who did travel < 24 km. Moreover, mothers who earn 1000-2000 (AOR= 3.10, 95%CI: 1.73-5.55) and > 2000 birr (AOR=2.66 95%CI: 1.52-4.64) had higher odds of maternal health service utilization than those who earn <500 birr. Similarly, the odds of utilizing maternal health service were higher among mothers who did not fear COVID-19 infection (AOR= 2.79, 95%CI: 1.85-4.20), who had not had to request permission from husband to visit the health facility (AOR= 7.24, 95%CI: 2.65-19.75), who had practiced COVID-19 prevention measure (AOR=5.82, 95%CI: 3.87-8.75), and used face mask (AOR= 2.06, 95% CI: 1.28-3.31) than their counterpart. Empowering mothers and creating awareness on the COVID-19 prevention is recommended to improve maternal health service utilization during the COVID-19 pandemic.

## Introduction

The novel coronavirus is a virus causing respiratory illness, commonly known as COVID-19 first noted in December 2019 in Wuhan, China, and has since then spread to countries in the world [1]. The World Health Organization (WHO) has declared the virus outbreak as a global pandemic on March 11, 2020 [2]. As of September 14, 2020, the virus has infected more than 28 million and killed more than 900,000 people [3]. Africa contributes 4% of infection (more than 1 million) and 3% deaths (more than 23,000) [3]. The WHO classifies maternal health services into: antenatal care (ANC), delivery services, and postnatal care, as essential health services to continue during the COVID-19 pandemics [4, 5].

The maternal deaths were predicted to rise by 17% in the best-scenario and 43% in the case of the worst scenario due to the COVID-19 pandemics [6]. Movement restrictions, transport challenges, and anxiety over possibly being exposed to coronavirus are acting as the barrier to maternal health service utilization [6, 7]. The ongoing COVID-19 has affected the perceptions of pregnant women; more than half of the participants in a study done in Naples hospital reported the psychological impact of COVID-19 as severe, and a similar number of pregnant women to worry of vertical transmission of the infection [7]. Strained health care systems, disruptions in care, and redirected resources might result in non-pandemic-related maternal morbidity and mortality and reproductive health crisis which are particularly true for African countries including Ethiopia with low-resource health systems [6, 8]. Obstetric healthcare providers may not be able to provide the highest quality care during the COVID-19 pandemic with essential precautions in place, client-provider communication is severely affected and the time to get the care needed may take longer as health workers try to protect themselves from the infection [9].

The International Confederation of Midwives and the Royal College of Midwives support community birth (home birth) for healthy women if there are appropriate midwife staffing and referrals are facilitated in obstetric emergencies, where these are not available, it may be necessary to modify available services, seeking at all times to maximize the provision of safe and positive birth experience to all women [4]. Ethiopia’s midwives grapple with COVID-19 while ensuring safe delivery, and maternal health workers reported that COVID-19 travel restrictions in Ethiopia are forcing pregnant women to give birth at home [8]. Maternity services in low resource countries are adapting to provide antenatal, delivery, and postnatal care amidst a rapidly shifting health system environment due to the COVID-19 pandemic [10]. Economic despair due to lost jobs, limited care, and restricted health services, overburdened health systems, restricted travel, and changing priorities at the primary care level are some of the burdens women had to face during the pandemic [11].

Many efforts have been taking place to enhance maternal health service utilization including information, education, and communication to raise awareness regarding the protection of mother and child during COVID-19, some countries have tried to open temporary birth centers, help hotlines virtual consultation with obstetricians have been provided via telemedicine services, to women seeking maternal health care service [6]. There is a consensus that the use of maternal health care services reduces maternal and child mortality and improves the reproductive health of women (12). Family planning, ANC, use of skilled delivery attendants, and PNC services are maternal health services that can significantly reduce maternal morbidity and mortality. Since the novel coronavirus (SARS-CoV-2) is new to humans, only limited scientific evidence is available to identify the impact of the disease on maternal health service delivery and utilization particularly in low resource settings such as Ethiopia. Accordingly, understanding the factors that affect maternal health care services delivery and utilization amid the COVID-19 pandemic could help to design appropriate strategies and policies towards the improvement of maternal health service provision and utilization. Thus, this study aimed to assess the maternal health care services (ANC, delivery, and PNC) utilization in the amid of the COVID-19 pandemic in West Shoa Zone, central Ethiopia.

## Materials and Methods

### Study design, setting, and population

A community-based cross-sectional study was conducted among women who were pregnant and those gave birth in the last 6 months of the study period in West Shoa Zone, central Ethiopia from July 1 to July 30, 2020. West Shoa zone is one of the zones of the Oromia National Regional state, its capital is located at114km distance to the western part of Addis Ababa, the capital city of Ethiopia. Currently, this zone has a total population of 2.5 million with around 1.2 million men and 1.3 women living within an area of 14,788.78 square kilometers. There are about 495,753 women of reproductive age and 100,283 mothers give births per year. West Shoa zone has 23 districts with over 528 rural kebele and 58 urban kebele. Currently, there are 8 Hospitals, 92 health centers, and 528 health posts in the West Shoa Zone.

### Sample size determination

The sample size was calculated using a single population proportion formula for the four maternal health services (ANC, Institutional delivery, and PNC). After calculating the sample size for the four maternal health services, the largest sample size was used as the overall sample size of the study. The assumptions considered during sample size calculation includes; 95% confidence interval, 5% the margin of error, and proportion of ANC visit 46% and proportion of institutional delivery 29% [13], and the proportion of early PNC visit 47% [14] in West Shoa zone from the previous study, and 10% non-response rate. The largest sample size calculated was for the proportion of early PNC visit 47%, n = 383, and used as the overall sample size of the study. The final sample size was 844 by considering 10 % non-response rate and the design effect of 2.

### Sampling techniques

A multi-stage stratified sampling technique was employed with strata of urban and rural kebele in the district. Six districts were selected at the first stage by simple random sampling from a total of 23 districts of West Shoa Zone. Then, kebele from the district was stratified into urban and rural kebele. Accordingly, one kebele from the urban and two kebeles from rural were enrolled from the selected districts by simple random sampling. Mothers who were pregnant or gave birth in the last 6 months of the study were identified by doing a census in collaboration with health extension workers through home to home visit. Then, number of the study population was placed to each selected kebele using proportion to population size. After having the list of all women in each selected kebele, mothers were included using systematic random sampling techniques every interval of four.

### Data collection tools and quality controls

Data were collected using semi-structured questionnaire through face-to-face interviews. The questionnaire was adapted from related literature and modified based on the objectives of the study and the cultural context of the study setting [3-8, 10, 11]. The tool consists of socio-demographic characteristics, maternal health care services amid COVID-19 pandemic (ANC, institutional delivery, and PNC) utilization, and impact of governmental emergency states (transport, distance, mask, and mandatory quarantine). Data collectors were 10 BSc health professionals, with health officers, Nurse, and Midwifery educational background. Pregnant mothers and those who gave birth in the last 6 months were identified first by using health extension workers to get their frame in the community. Data quality was assured during collection, coding, entry, and analysis. A two-days training was given for the data collectors and supervisors before actual data collection. The collected data were reviewed and checked for consistency, clarity; completeness, and accuracy throughout the data collection process by data collectors and supervisors. Furthermore, the pretest was done among 5% of the sample and the result was used to modify the tool.

### Data processing and analysis

Collected data were cleaned and entered using Epi data version 4.6 and analyzed using SPSS version 23. Descriptive statistics were employed using frequencies, percentages, and diagrams. Both bivariable and multivariable logistic regressions were performed to identify the presence of a significant association between independent variables and the dependent variable. Those variables with p<0.2 in the bivariable analysis were considered for multivariable logistic regression analysis. Hosmer and Lemeshow goodness of fit test was used to check model fitness before running the final model. Finally, screened variables were fitted to the multivariable logistic regression model through a backward stepwise method to reduce the effects of cofounders and to identify the independent effects of each variable on the outcome variable. An adjusted odds ratio for a 95% confidence interval was employed for the strength and directions of association between independent variables and the outcome variables. A P-value of <0.05 was used to declare statistical significance.

### Operational Definitions

Maternal health service utilization was defined as; fully received ANC in line with their gestational age, having institutional delivery, and any PNC visits during COVID-19. Those maternal health care services utilization were assessed using yes or no questions.

### Ethics statement

The ethical clearance was obtained from an ethical review committee of the college of medicine and health sciences, Ambo University. A letter of support was submitted to each district health office. Both written and verbal consent was obtained from each study subject before the data collection process proceeded. During the data collection process, the data collectors had informed each study participant about the objective and anticipated benefits of the research project and the study participants were also informed of their full right to refuse, withdraw, or completely reject part or all of their parts in the study.

## Result

### Socio-demographic and economic characteristics of the study participants

In this study, a total of 844 pregnant women and those who gave birth in the last 6 months were included; the response rate was 100%. Two hundred forty-two participants (28.7%) were between the age of 25-29 years and 320 (37.9%) of they had primary education. More than half, 467 (55.3%) of the participants’ occupations were housewives (Table 1).

**Table 1:** Sociodemographic distribution of participants showing maternal health services in the amid of the COVID-19 pandemic in West Shoa Zone, Central Ethiopia, 2020 (n= 844)

| Characteristics | Frequency | Percentage |
| --- | --- | --- |
| **Maternal age** |  |  |
| < 19 years | 43 | 5.1 |
| 20-24 years | 229 | 27.1 |
| 25-29 years | 242 | 28.7 |
| 30-34 years | 198 | 23.5 |
| 35-39 years | 102 | 12.1 |
| ≥40 years | 30 | 3.6 |
| **Marital status** |  |  |
| Married | 812 | 96.2 |
| Others* | 32 | 3.8 |
| **Maternal education** |  |  |
| Cannot read and write | 165 | 19.5 |
| Primary education | 320 | 37.9 |
| Secondary education | 243 | 28.8 |
| college and above | 116 | 13.7 |
| **Place of Residence** |  |  |
| Urban | 560 | 66.4 |
| Rural | 284 | 33.6 |
| **Estimated monthly income in birr** |  |  |
| < 500 | 159 | 18.8 |
| 500-1000 | 200 | 23.7 |
| 1000-2000 | 182 | 21.6 |
| > 2000 | 303 | 35.9 |
| **Maternal occupation** |  |  |
| Housewife | 467 | 55.3 |
| Government employee | 117 | 13.9 |
| Self-employed | 146 | 17.3 |
| Student | 51 | 6.0 |
| Farmer | 63 | 7.5 |
| **Husband occupation** |  |  |
| Government employee | 222 | 27.5 |
| Self-employed | 291 | 36.1 |
| Student | 19 | 2.4 |
| Farmer | 275 | 34.1 |
| **Distance to the health facility** |  |  |
| ≤24 km | 531 | 62.9 |
| 25-74 km | 168 | 19.9 |
| 75-99 km | 115 | 13.6 |
| ≥100 km | 30 | 3.6 |

### Obstetric characteristics of the study participants

Among the 844 participants, 329(39.0%) were prim gravid and 657(77.8%) of them had 1-5 deliveries (parity) (Table 2).

**Table 2:** Obstetric distribution of study participants to assess maternal health services utilization in the amid of the COVID-19 pandemic in West Shoa Zone, Central Ethiopia, 2020 (n= 844)

| Characteristics | Frequency | Percentage |
| --- | --- | --- |
| **Previous adverse pregnancy outcome** |  |  |
| Yes | 165 | 19.5 |
| No | 679 | 80.5 |
| **Gravidity** |  |  |
| Prim gravida | 329 | 39.0 |
| 2-4 | 468 | 55.5 |
| ≥5 | 47 | 5.6 |
| **Parity** |  |  |
| Null parity | 165 | 19.5 |
| 1-4 | 657 | 77.8 |
| ≥5 | 22 | 2.6 |

### Awareness on COVID-19 modes of transmission and prevention mechanisms

Among 844 of the participants, 835(98.9%) of them responded that COVID-19 can transmit from person to person and 230(27.3%) of them think they are at risk of getting the COVID-19 infection. Five hundred fifty-three, 65.5% were practicing COVID-19 prevention measures that they had received from the community (Table 3).

**Table 3:** COVID-19 related characteristics to assess maternal health service utilization among pregnant women and gave birth in the last 6 months of the study period in West Shoa Zone, Central Ethiopia, 2020 (n= 844)

| Characteristics | Frequency | Percentage |
| --- | --- | --- |
| **COVID-19 can transmit from person to person** |  |  |
| Yes | 835 | 98.9 |
| No | 9 | 1.1 |
| **Handshaking with a person having the virus can transmit COVID-19** |  |  |
| Yes | 812 | 96.2 |
| No | 32 | 3.8 |
| **Sitting nearby a person having the virus can transmit COVID-19** |  |  |
| Yes | 554 | 65.6 |
| No | 290 | 34.4 |
| **Body fluid of a person having the virus can transmit COVID-19** |  |  |
| Yes | 294 | 34.8 |
| No | 550 | 65.2 |
| **Cough of a person having the virus can transmit COVID-19** |  |  |
| Yes | 639 | 75.7 |
| No | 205 | 24.3 |
| **Sexual intercourse with an infected person can transmit COVID-19** |  |  |
| Yes | 25 | 3.0 |
| No | 819 | 97.0 |
| **At the risk of getting the COVID-19** |  |  |
| Yes | 230 | 27.3 |
| No | 614 | 72.7 |
| **Confidence to protect from COVID-19** |  |  |
| Yes | 763 | 90.4 |
| No | 81 | 9.6 |
| **Fear to visit the health facility** |  |  |
| Yes | 481 | 57 |
| No | 363 | 43 |
| **Information about COVID-19 prevention** |  |  |
| Yes | 579 | 68.6 |
| No | 265 | 31.4 |
| **Has sanitizer and/or alcohol** |  |  |
| Yes | 191 | 22.6 |
| No | 653 | 77.4 |
| **Use of face mask** |  |  |
| Yes | 609 | 77.2 |
| No | 235 | 27.8 |
| **Practicing COVID-19 prevention measures** |  |  |
| Yes | 553 | 65.5 |
| No | 291 | 34.5 |
| **Request permission from the husband to visit the health facility (n=812)** |  |  |
| Yes | 735 | 90.5 |
| No | 77 | 9.5 |
| **Media presence** |  |  |
| Yes | 722 | 85.5 |
| No | 122 | 14.5 |

### Reasons for not utilizing maternal health services

Among the 297 participants who did not utilize maternal health services, 109(36.5%) mention fear of getting COVID-19 while traveling to receive the service as a reason to not receive the service, and 96(32.3%) and 95 (32%) gave fear of COVID-19 transmission from health care providers and lack of sanitizer or water in a health facility as a reason not to utilize the maternal health services respectively (Fig 1).

**Fig 1.** Reasons for not using maternal health service utilization during COVID-19, West Shoa Zone, central Ethiopia, 2020

### Prevalence of maternal health service utilization and associated factors

In this study, about two-thirds of 547(64.8%) [95% CI: 61.5-68.0] of mothers received maternal health care service during COVID-19. Binary logistic regression was fitted to assess the statistical association between factors and maternal health service utilization during the COVID-19 pandemic. All factors with p-value ≤0.2 on bi-variable logistic regression were taken to multivariable analysis to control for potential confounding effects. Accordingly, maternal age, maternal occupation, husband occupation, educational status, place of residence, distance from health the facility, estimated monthly income, media presence, marital status, gravidity, parity, face mask use, fear of COVID-19 infections, permission request from husband to visit a health facility, sitting nearby a person having the virus can transmit COVID-19, cough of a person having the virus can transmit COVID-19, sexual intercourse with an infected person can transmit COVID-19, risk of COVID-19 infection, confidence to protect COVID-19, received information about COVID-19 prevention, the practice of COVID-19 prevention measure had a p-value < 0.2 at bi-variable logistic regression and therefore taken to the multi-variable logistic regression. However, maternal educational status, distance from the health facility, monthly estimated income, fear of COVID-19 infection, permission request from husband to visit a health facility, and practicing COVID-19 prevention measures were significantly associated with maternal health service utilization at multivariable logistic regression.

The odds of maternal health service utilization during the COVID-19 pandemic in mothers who had primary education were 2.16 times higher than those who could not read and write (AOR=2.16, 95% CI: 1.29-3.60). Similarly, mothers who had secondary education had higher odds of maternal health service utilization during the COVID-19 pandemic than those who could not read and write (AOR=1.97, 95% CI: 1.13-3.44). Moreover, the odds of maternal health service utilization during the COVID-19 pandemic in mothers who had college and above education were 2.89 times higher than those who could not read and write (AOR=2.89, 95% CI:1.34-6.22).

Mothers who did travel 25-74 km to reach health facility were 63% less likely to utilize maternal health service during the COVID-19 pandemic than those who did travel < 24 km to reach health facility (AOR= 0.37, 95% CI:0.23-0.59). Furthermore, mothers who had to travel 75-99 km to reach health facilities were 90% less likely to utilize maternal health service during the COVID-19 pandemic than those who did travel < 24 km to reach health facility (AOR= 0.10, 95% CI:0.05-0.19). While, mothers who earn 1000-2000 birr had higher odds of utilizing maternal health service during the COVID-19 pandemic than those who earn <500 birr (AOR= 3.10, 95% CI: 1.73-5.55). Likewise, the odds of maternal health service utilization during the COVID-19 pandemic were 2.66 higher in mothers who earn ≥2000 birr than those who earn <500 birr (AOR=2.66, 95% CI: 1.52-4.64).

The odds of utilizing maternal health care service during the COVID-19 pandemic in mothers who did not fear COVID-19 infection was 2.79 times higher than in those who did fear COVID-19 infection (AOR= 2.79, 95% CI:1.85-4.20). Moreover, mothers who had not had to request permission from husband to visit the health facility had 7.24 times higher odds of utilizing maternal health care service during the COVID-19 pandemic than those who had to request permission from husband to visit the health facility (AOR= 7.24, 95% CI: 2.65-19.75). The odds of utilizing maternal health care services during the COVID-19 pandemic in those who practice COVID-19 prevention measure were 5.82 times higher than in those who did not practice the prevention measures (AOR=5.82, 95% CI: 3.87-8.75). The odds of maternal health services utilization amidst COVID-19 were 2.06 times higher among mothers who use face mask than those mothers who did not use a face mask (AOR= 2.06, 95% CI:1.28-3.31) (Table 4).

**Table 4:**
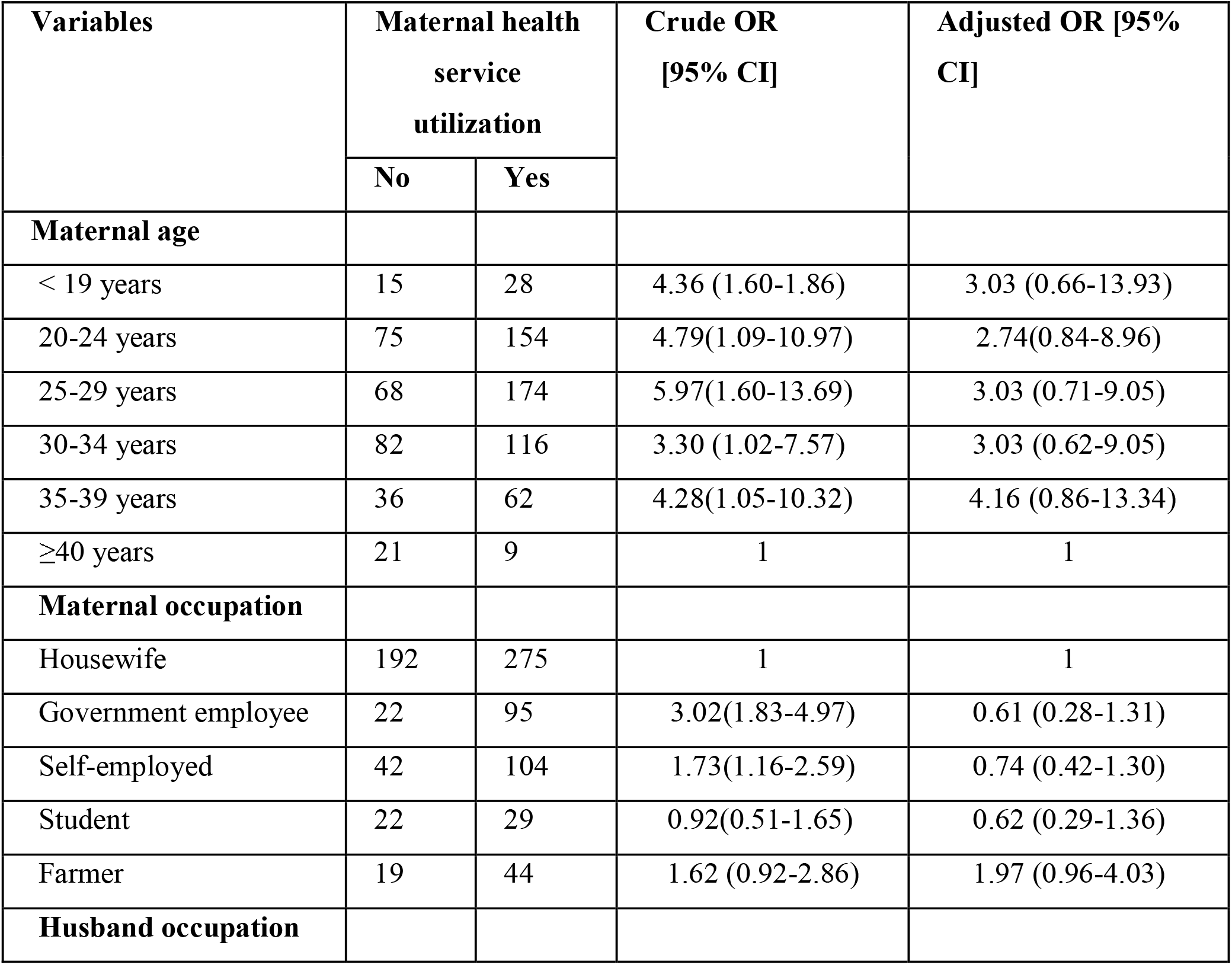
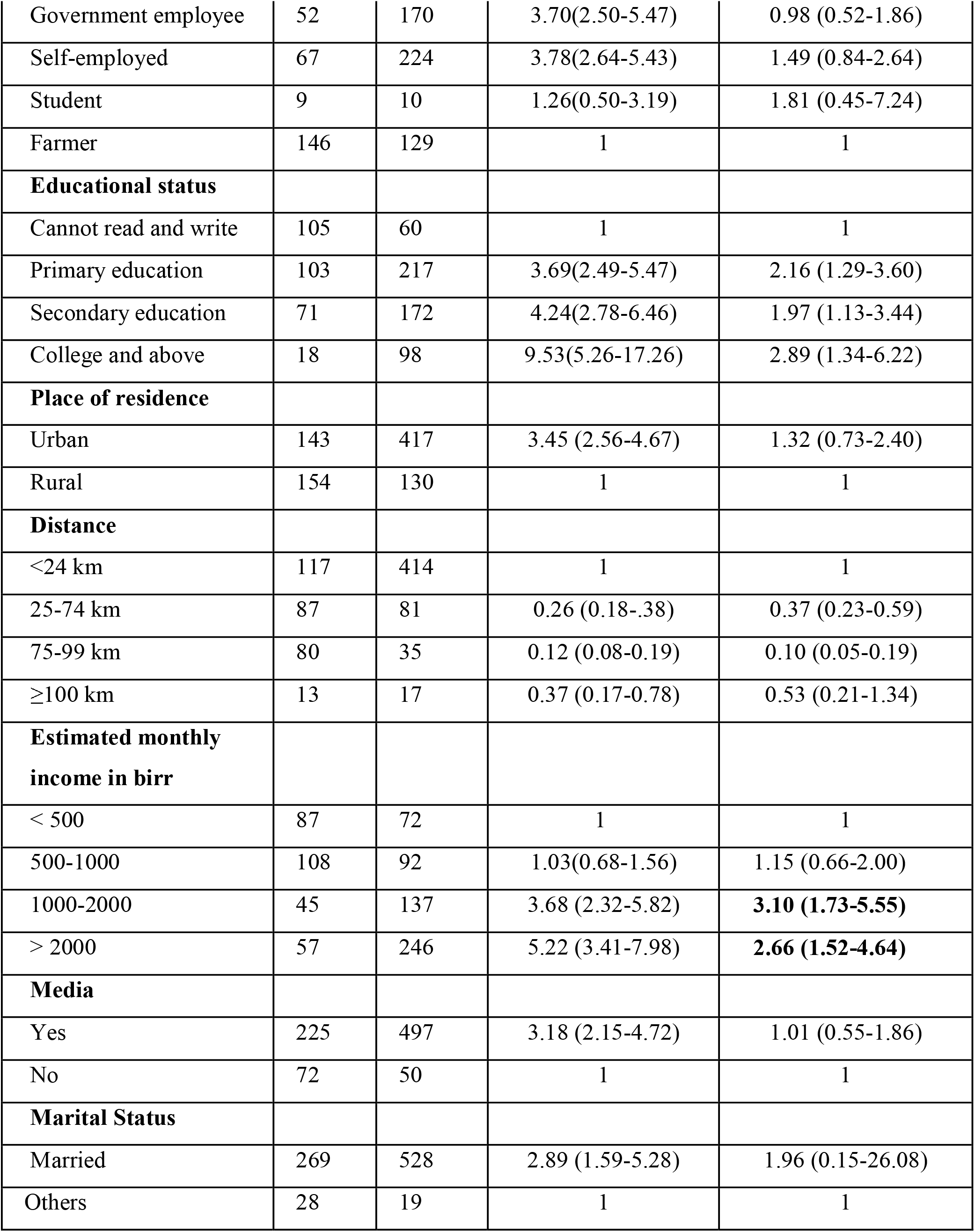
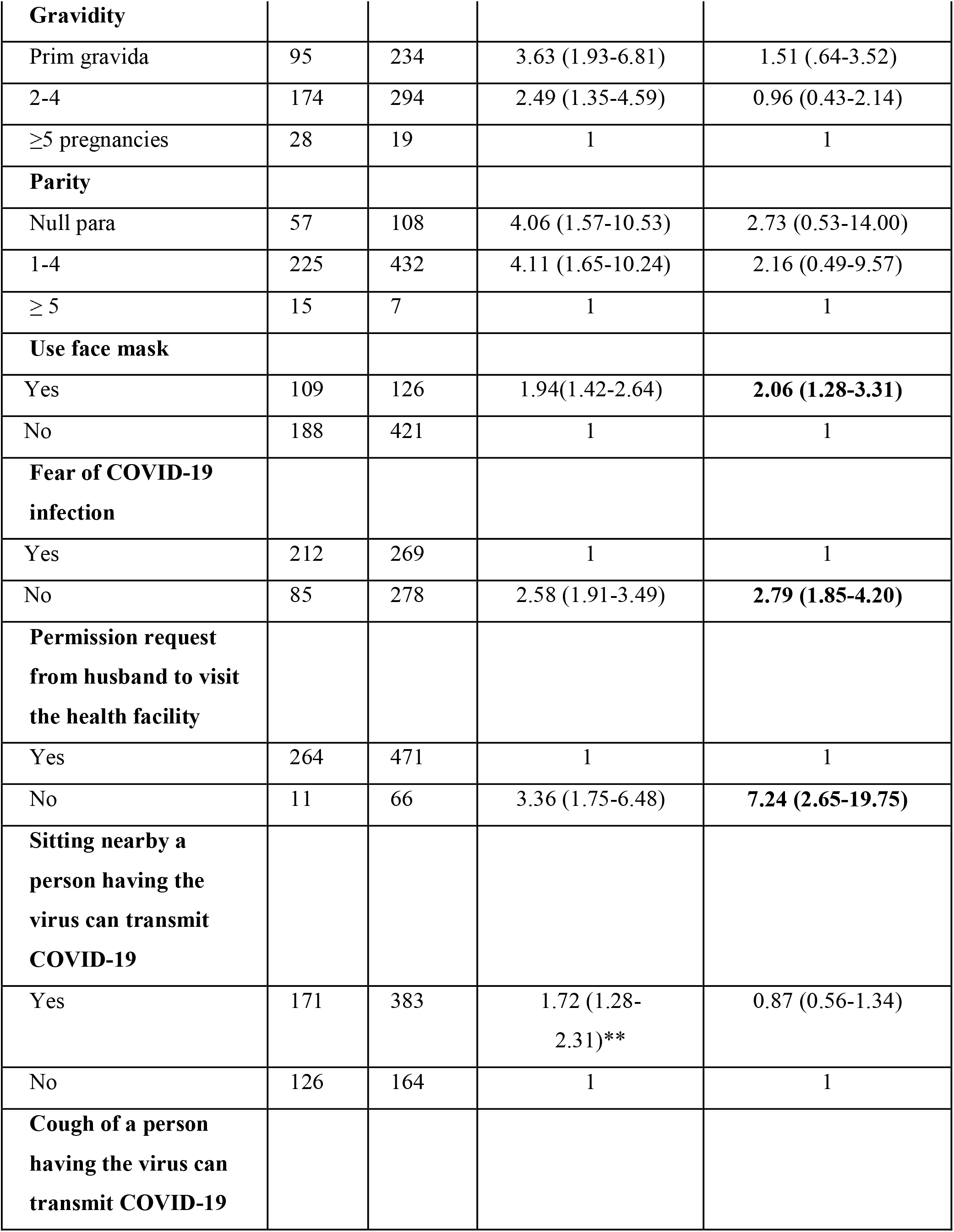
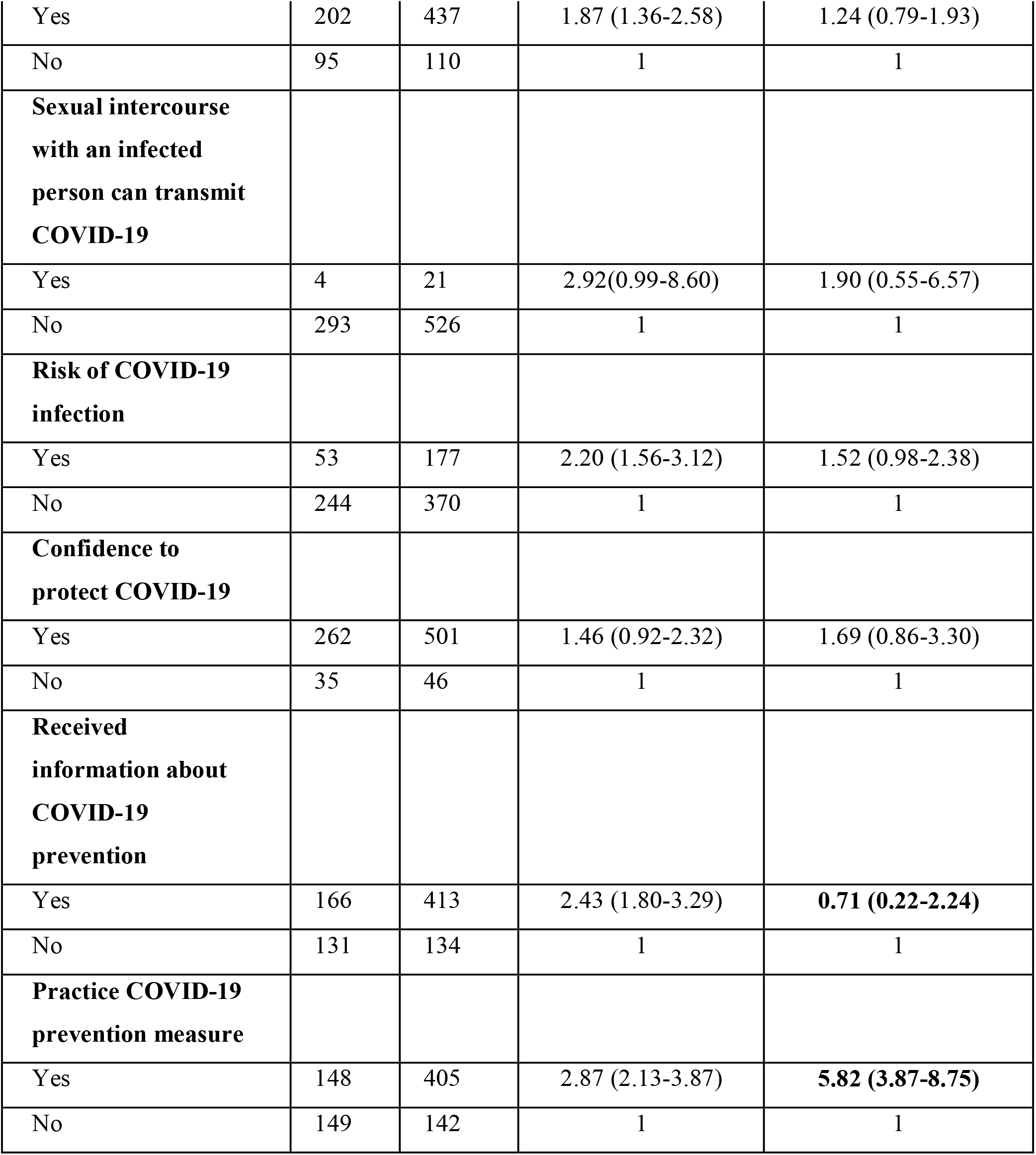
Bi-variable and multi-variable logistic regression to assess factors associated with maternal health services in the amid of the COVID-19, West Shoa Zone, central Ethiopia, 2020 (n=844)

| Variables | Maternal health service utilization |  | Crude OR [95% CI] | Adjusted OR [95% CI] |
| --- | --- | --- | --- | --- |
|  | No | Yes |  |  |
| **Maternal age** |  |  |  |  |
| < 19 years | 15 | 28 | 4.36 (1.60-1.86) | 3.03 (0.66-13.93) |
| 20-24 years | 75 | 154 | 4.79(1.09-10.97) | 2.74(0.84-8.96) |
| 25-29 years | 68 | 174 | 5.97(1.60-13.69) | 3.03 (0.71-9.05) |
| 30-34 years | 82 | 116 | 3.30 (1.02-7.57) | 3.03 (0.62-9.05) |
| 35-39 years | 36 | 62 | 4.28(1.05-10.32) | 4.16 (0.86-13.34) |
| ≥40 years | 21 | 9 | 1 | 1 |
| **Maternal occupation** |  |  |  |  |
| Housewife | 192 | 275 | 1 | 1 |
| Government employee | 22 | 95 | 3.02(1.83-4.97) | 0.61 (0.28-1.31) |
| Self-employed | 42 | 104 | 1.73(1.16-2.59) | 0.74 (0.42-1.30) |
| Student | 22 | 29 | 0.92(0.51-1.65) | 0.62 (0.29-1.36) |
| Farmer | 19 | 44 | 1.62 (0.92-2.86) | 1.97 (0.96-4.03) |
| **Husband occupation** |  |  |  |  |
| Government employee | 52 | 170 | 3.70(2.50-5.47) | 0.98 (0.52-1.86) |
| Self-employed | 67 | 224 | 3.78(2.64-5.43) | 1.49 (0.84-2.64) |
| Student | 9 | 10 | 1.26(0.50-3.19) | 1.81 (0.45-7.24) |
| Farmer | 146 | 129 | 1 | 1 |
| **Educational status** |  |  |  |  |
| Cannot read and write | 105 | 60 | 1 | 1 |
| Primary education | 103 | 217 | 3.69(2.49-5.47) | 2.16 (1.29-3.60) |
| Secondary education | 71 | 172 | 4.24(2.78-6.46) | 1.97 (1.13-3.44) |
| College and above | 18 | 98 | 9.53(5.26-17.26) | 2.89 (1.34-6.22) |
| **Place of residence** |  |  |  |  |
| Urban | 143 | 417 | 3.45 (2.56-4.67) | 1.32 (0.73-2.40) |
| Rural | 154 | 130 | 1 | 1 |
| **Distance** |  |  |  |  |
| <24 km | 117 | 414 | 1 | 1 |
| 25-74 km | 87 | 81 | 0.26 (0.18-.38) | 0.37 (0.23-0.59) |
| 75-99 km | 80 | 35 | 0.12 (0.08-0.19) | 0.10 (0.05-0.19) |
| ≥100 km | 13 | 17 | 0.37 (0.17-0.78) | 0.53 (0.21-1.34) |
| **Estimated monthly income in birr** |  |  |  |  |
| < 500 | 87 | 72 | 1 | 1 |
| 500-1000 | 108 | 92 | 1.03(0.68-1.56) | 1.15 (0.66-2.00) |
| 1000-2000 | 45 | 137 | 3.68 (2.32-5.82) | **3.10 (1.73-5.55)** |
| > 2000 | 57 | 246 | 5.22 (3.41-7.98) | **2.66 (1.52-4.64)** |
| **Media** |  |  |  |  |
| Yes | 225 | 497 | 3.18 (2.15-4.72) | 1.01 (0.55-1.86) |
| No | 72 | 50 | 1 | 1 |
| **Marital Status** |  |  |  |  |
| Married | 269 | 528 | 2.89 (1.59-5.28) | 1.96 (0.15-26.08) |
| Others | 28 | 19 | 1 | 1 |
| **Gravidity** |  |  |  |  |
| Prim gravida | 95 | 234 | 3.63 (1.93-6.81) | 1.51 (.64-3.52) |
| 2-4 | 174 | 294 | 2.49 (1.35-4.59) | 0.96 (0.43-2.14) |
| ≥5 pregnancies | 28 | 19 | 1 | 1 |
| **Parity** |  |  |  |  |
| Null para | 57 | 108 | 4.06 (1.57-10.53) | 2.73 (0.53-14.00) |
| 1-4 | 225 | 432 | 4.11 (1.65-10.24) | 2.16 (0.49-9.57) |
| ≥ 5 | 15 | 7 | 1 | 1 |
| **Use face mask** |  |  |  |  |
| Yes | 109 | 126 | 1.94(1.42-2.64) | **2.06 (1.28-3.31)** |
| No | 188 | 421 | 1 | 1 |
| **Fear of COVID-19 infection** |  |  |  |  |
| Yes | 212 | 269 | 1 | 1 |
| No | 85 | 278 | 2.58 (1.91-3.49) | **2.79 (1.85-4.20)** |
| **Permission request from husband to visit the health facility** |  |  |  |  |
| Yes | 264 | 471 | 1 | 1 |
| No | 11 | 66 | 3.36 (1.75-6.48) | **7.24 (2.65-19.75)** |
| **Sitting nearby a person having the virus can transmit COVID-19** |  |  |  |  |
| Yes | 171 | 383 | 1.72 (1.28-2.31)** | 0.87 (0.56-1.34) |
| No | 126 | 164 | 1 | 1 |
| **Cough of a person having the virus can transmit COVID-19** |  |  |  |  |
| Yes | 202 | 437 | 1.87 (1.36-2.58) | 1.24 (0.79-1.93) |
| No | 95 | 110 | 1 | 1 |
| **Sexual intercourse with an infected person can transmit COVID-19** |  |  |  |  |
| Yes | 4 | 21 | 2.92(0.99-8.60) | 1.90 (0.55-6.57) |
| No | 293 | 526 | 1 | 1 |
| **Risk of COVID-19 infection** |  |  |  |  |
| Yes | 53 | 177 | 2.20 (1.56-3.12) | 1.52 (0.98-2.38) |
| No | 244 | 370 | 1 | 1 |
| **Confidence to protect COVID-19** |  |  |  |  |
| Yes | 262 | 501 | 1.46 (0.92-2.32) | 1.69 (0.86-3.30) |
| No | 35 | 46 | 1 | 1 |
| **Received information about COVID-19 prevention** |  |  |  |  |
| Yes | 166 | 413 | 2.43 (1.80-3.29) | **0.71 (0.22-2.24)** |
| No | 131 | 134 | 1 | 1 |
| **Practice COVID-19 prevention measure** |  |  |  |  |
| Yes | 148 | 405 | 2.87 (2.13-3.87) | **5.82 (3.87-8.75)** |
| No | 149 | 142 | 1 | 1 |

## Discussion

This study found the prevalence of maternal health service utilization was 64.8% [95%CI: 61.5-68.0]. This prevalence is lower than the studies done in Ethiopia before the COVID-19 pandemic [14, 15]. This might be due to the lockdown and movement restrictions brought about by the ongoing pandemic and lack of transportation manifested as a result [16]. Also, the dread and anxiety of visiting hospitals during COVID-19 has led many women to change their plan of childbirth and they are planning to have home deliveries [6].

The current study reported that the odds of maternal health service utilization during the COVID-19 pandemic were higher among mothers who had primary, secondary, and college and above education than those who could not read and write. This is consistent with the study done in developing countries [17], and Nigeria [18]. Moreover, mothers who earn more than 1000 birr per month had higher odds of utilizing maternal health service during the COVID-19 pandemic than those who earn < 500 birr. This is in agreement with the study done in developing countries [17], and Nepal [19]. This is because economically independent women do not rely on other people to decide to utilize maternal health services [20].

Furthermore, mothers who did travel 25-74 km and 75-99 km to reach the health facility were 63% and 90% less likely to utilize maternal health service during the COVID-19 pandemic than those who did travel < 24 km to reach the health facility respectively. This might be due to a change of plan of ANC follow-up, childbirth, and other maternal health services hence the first delay, and planning to have home deliveries [6, 20]. Besides, the odds of utilizing maternal health service during the COVID-19 pandemic in mothers who did not fear COVID-19 infection was 2.79 times higher than those who did fear COVID-19 infection. This could be due to the psychological and social effects of the COVID-19 pandemic which is more pronounced in women forcing them to plan home deliveries not to expose their family to the infection [6]. Besides, mothers who had not had to request permission from husband to visit health facility had the higher odds of utilizing maternal health service during the COVID-19 pandemic than those who had to request permission from the husband to visit a health facility. This is consistent with the studies done in Ethiopia [14, 21]. This might be due to women’s autonomy is positively associated with maternal health service utilization. Also, women with control over resources including physical, human, intellectual, and financial resources are more independent in deciding and seeking health care [21]. The odds of utilizing maternal health service during the COVID-19 pandemic in those who practice COVID-19 prevention measure was 5.82 times higher than in those who did not practice the prevention measure Similarly, the odds of maternal health service utilization amidst COVID-19 were 2.06 times in mothers who use face mask than those mothers who did not use a face mask. This is following the study done in Iran [22].

## Conclusion

In this study, the prevalence of maternal health service utilization was found low. Furthermore, maternal educational status, distance from the health facility, monthly estimated income, fear of COVID-19 infection, permission request from husband to visit a health facility, and practicing COVID-19 prevention measures were found to be significantly associated with maternal health service utilization. The federal ministry of health and regional health bureau should work in collaboration to aware of the community and mothers, in particular, to increase the maternal health service during the pandemic while practicing the necessary preventive measures to control the spread of the infection.

## Data Availability

All the datasets used and analyzed during the current study are available from the manuscript.

## List of abbreviations

ANC: Antenatal Care
COVID□19: Corona Virus Disease 2019
PNC: Postnatal Care
WHO: World Health Organization

## Acknowledgments

The authors are grateful to the participants of the study who shared their time to give their genuine responses, data collectors, and supervisors of the study.

