## Supplemental Figure 1 for "Maternal health care services utilization in the amid of COVID-19 pandemic in West Shoa Zone, Central Ethiopia"

Reasons for not utilizing maternal health services

|  | yes | No | #REF! |
| --- | --- | --- | --- |
| Fear of COVID-19 transmission from health care providers | 96 | 201 | 1 |
| Fear of getting COVID-19 while receiving the services | 109 | 188 |  |
| Fear of getting COVID-19 while travelling to receive the service | 62 | 235 |  |
| Increment of tariff of transportation | 59 | 238 |  |
| service providers are not giving the service as they were before | 90 | 207 |  |
| Examination materials are not clean | 64 | 233 |  |
| Long waiting hours and over crowdedness | 61 | 236 |  |
| Lack of sanitizer or water in health facility | 95 | 202 |  |
| Health care provider has no personel protective equipments | 64 | 233 |  |
